# NeuroAid: An Open-Data Multimodal Screening Framework for Parkinson’s and Depression Risk Estimation

**DOI:** 10.64898/2026.08.03.26359539

**Authors:** R L Jayesh, U Rajath, Prachi Patel

**Affiliations:** Dept. of CSE (AI & ML) Dayananda Sagar University Bengaluru, India

**Keywords:** Parkinson’s disease screening, depression risk estimation, multimodal fusion, clinical AI, open-data benchmarking, speech foundation models, participant-safe splitting, class-balanced evaluation, MLOps, digital phenotyping

## Abstract

Neurological and mental-health conditions such as Parkinson’s disease (PD) and major depressive disorder (MDD) impose a substantial and growing global burden, yet reliable early screening remains largely confined to specialist clinical settings that are inaccessible to the majority of affected individuals. We present NeuroAid, an open-data multimodal AI screening framework that estimates condition-specific risk from non-invasive, accessible signals spanning acoustic speech biomarkers, facial and video-based affective cues, and clinical or behavioral digital biomarkers. NeuroAid is organized as a modular, branch-wise pipeline covering three independent signal pathways—audio, vision, and behavioral—unified by a frozen-embedding late-fusion layer that produces interpretable joint risk scores. A participant-safe, subject-grouped splitting protocol is enforced throughout, preventing inter-subject data leakage—a frequently overlooked cause of artificially inflated performance in clinical machine learning benchmarks. On the Figshare Parkinson’s audio dataset, a proposed small-data protocol combining frozen WavLM foundation-model embeddings with a grouped SVM-RBF classifier achieves a cross-validated balanced accuracy of 0.786±0.073 and a held-out test balanced accuracy of 75.0%, F1-score of 80.0%, and AUC-ROC of 82.8%. The depression vision branch, trained on the DepVidMood corpus via transfer from FER-2013, reaches a threshold-tuned test balanced accuracy of 59.7% and is presented as an honest hard-case baseline under severe class imbalance. NeuroAid is further distinguished by its production-grade MLOps scaffolding: orchestrated branch training, JSON and Markdown artifact reporting, a deployable Streamlit screening interface, and a complete CI/CD workflow. The entire system is built exclusively on publicly available datasets, ensuring full reproducibility. All code, artifacts, and benchmark outputs are versioned and deployable via Docker.

## I. Introduction

Neurological and psychiatric disorders represent two of the most rapidly escalating and chronically underfunded challenges in global healthcare. Parkinson’s disease affects an estimated 10 million individuals worldwide [1], [16], and its global prevalence is projected to double by 2040 as populations age. Critically, the dopaminergic neuronal degeneration underlying PD begins a decade or more before the onset of classic motor symptoms such as resting tremor and rigidity [3]. During this prodromal window, hypokinetic dysarthria—a motor speech disorder characterized by reduced vocal intensity, flattened pitch inflection, and imprecise articulation—is among the earliest observable manifestations, present in up to 89% of PD patients at initial assessment and detectable from sustained phonation recordings. Major depressive disorder affects more than 300 million people globally and is projected among the leading causes of disability burden [15], [17]. Depression is similarly characterized by measurable pre-clinical behavioral signatures: psychomotor slowing, affective flattening, reduced facial action variability, and altered speech prosody can all be captured from video and audio recordings well before a formal clinical diagnosis is established [7], [20].

Despite the existence of these early-detectable signal signatures, the current clinical pathway for both conditions remains reactive rather than proactive. Parkinson’s is typically diagnosed via the Movement Disorder Society Unified PD Rating Scale (MDS-UPDRS) or DaT-SPECT neuroimaging [18]— both requiring specialist access. Depression diagnosis relies on structured psychiatric interviews (PHQ-9, HAM-D) that are administered episodically and subject to substantial self-report bias. These barriers are compounded by a shortage of neurologists and psychiatrists in primary-care and resource-limited settings, creating a diagnostic gap that disproportionately affects populations least able to seek specialist care.

The machine learning literature has proposed numerous systems for automated screening from audio, video, and tabular clinical signals. However, this body of work exhibits three systematic weaknesses that limit its real-world impact [19]. First, the majority of high-performing systems rely on proprietary or restricted-access datasets that cannot be replicated by independent groups [5]. Second, evaluation protocols frequently allow recording-level data leakage—where multiple recordings from the same participant appear in both training and test partitions—producing performance estimates that are optimistic by ten to twenty percentage points [11]. Third, research pipelines rarely propagate model improvements through to deployable inference artifacts, creating a persistent disconnect between benchmark results and usable screening tools. These weaknesses are especially damaging for clinical AI systems, where reproducibility and calibrated uncertainty are prerequisites for safe deployment [14].

NeuroAid is designed to address all three gaps within a single, unified open-data framework. It adopts a strictly open-data philosophy, building every model on publicly available datasets. It enforces subject-aware evaluation throughout— including participant-grouped cross-validation and threshold tuning optimized for balanced accuracy—to produce honest, leakage-free performance estimates. And it integrates research-grade benchmarking with a production-oriented MLOps stack such that training gains always propagate to inference artifacts, downloadable reports, a live Streamlit demo, and a Docker-deployable container. The system is explicitly positioned not as a clinical diagnosis tool, but as a reproducible, deployable open-data screening research platform whose primary contribution is the integration of heterogeneous clinical signals into a single auditable, honestly evaluated research-to-demo workflow.

The principal contributions of this work are:

- A modular multimodal screening architecture unifying audio, vision, and behavioral signal branches under a frozen-embedding late-fusion layer, trained exclusively on open, publicly available datasets.
- A small-data protocol for Parkinson’s audio screening combining frozen WavLM embeddings with participant-grouped SVM-RBF classification that achieves balanced accuracy of 75.0%, F1 of 80.0%, and AUC of 82.8% on held-out participants—a 23-point improvement over naive end-to-end fine-tuning.
- A threshold-aware, class-balanced benchmarking framework for depression vision screening that presents an honest hard-case baseline (59.7% balanced accuracy after threshold tuning) rather than masking class-imbalance failures behind inflated raw accuracy.
- An end-to-end MLOps pipeline including orchestrated branch training, per-stage skip/fail semantics, machine-readable JSON artifact reporting, GitHub Actions CI/CD workflows, and a Streamlit demonstration application with downloadable benchmark reports.
- A rigorous ablation demonstrating that recording-level splitting inflates Parkinson’s audio balanced accuracy by 16 percentage points relative to participant-safe evaluation, quantifying the cost of a pervasive methodological failure in clinical ML.

## II. Literature Review

### A. Acoustic Biomarkers for Parkinson’s Disease

The use of acoustic features for PD screening has a history spanning more than two decades. Little *et al*. [6] established the foundational result that dysphonia measures derived from sustained phonation— including RPDE (recurrence period density entropy) and DFA (detrended fluctuation analysis)— could discriminate PD from healthy controls with accuracy exceeding 91% on the UCI Parkinson’s dataset. Subsequent work broadened the feature space to include Mel-frequency cepstral coefficients, jitter, shimmer, and harmonics-to-noise ratio, and applied classical classifiers (SVM, random forest, AdaBoost) and shallow neural networks in increasingly elaborate ensembles.

The transition to deep learning introduced spectrogram-based approaches. Tekindor and Aydin [4] proposed a hybrid convolutional autoencoder with LSTM sequence modeling on sustained phonation vowels, achieving 95.79% accuracy on a specific scripted corpus. While high, this result is reported under recording-level evaluation where multiple recordings from the same speaker may leak across train and test partitions—an evaluation setting the authors do not explicitly address.

The most significant recent development is the application of pre-trained speech foundation models. Brueckner *et al*. [3] introduced a new in-clinic free-speech PD corpus and demonstrated that frozen HuBERT and Wav2Vec2 embeddings achieve unweighted average recall (UAR) of approximately 0.97 when combined with lightweight downstream classifiers. Their results establish that large speech models pretrained on hundreds of thousands of hours of general speech capture representations that transfer directly to pathological voice classification, even from brief utterances. The Trillsson small-model variant further shows that on-device screening is computationally feasible. NeuroAid adopts WavLM as its foundation speech encoder following this paradigm, extending it with a participant-grouped evaluation protocol that Brueckner *et al*. do not employ. Puerta-Acevedo *et al*. [5] survey ten publicly available PD speech corpora and document the inconsistency of demographic coverage, recording tasks, and annotation quality across these resources, providing the principled justification for NeuroAid’s decision to use the Figshare PD corpus as its primary audio evaluation benchmark.

Beyond foundation models, recent work has extended PD audio analysis to novel mathematical representations. Hybrid spectral, topological, and random matrix methods [21] leverage persistent homology to capture non-linear vocal dynamics that MFCC-based features overlook, achieving a peak accuracy of 98.0% and F1 of 97.98% with an MLP classifier trained on synthetically augmented speech. Explainability has also emerged as a priority: an attention-augmented ResNet with Squeeze-and-Excitation blocks [24] employs Grad-CAM, integrated gradients, and LIME to identify which temporal regions of voice recordings carry pathological dysphonic information, achieving 97.8% accuracy while providing clinician-readable justifications. NeuroAid identifies XAI for the audio branch as a future roadmap item, recognizing that clinician-facing deployment requires interpretable explanations alongside risk scores.

### B. Facial and Video-Based Depression Screening

Facial action coding and video-based affective analysis have been extensively explored as non-invasive proxies for depression severity. A large body of work extracts action unit (AU) intensities using OpenFace or similar pipelines and feeds these into SVMs, random forests, or regression models for PHQ-8 score prediction. The AVEC challenge series [9] has served as the primary benchmark environment for this line of work, though access to AVEC data is restricted, limiting reproducibility.

More recent approaches leverage deep transfer learning. Lee *et al*. [7] introduced the use of inter-subject correlation (ISC) of AU time series as a novel feature, demonstrating 72–90% classification accuracy on a controlled sample of 98 participants. Their result is notable because it captures the dynamic temporal alignment of facial expression across individuals—a signal that static frame-level classifiers miss entirely. Dalal *et al*. [8] take a complementary approach, incorporating PHQ-9 clinical guidelines into a cross-attention architecture to produce depression predictions that align with the diagnostic criteria clinicians actually use, addressing the interpretability barrier that limits adoption of purely data-driven models. A persistent challenge across all depression vision work is severe class imbalance: healthy and neutral facial states dominate most naturalistic video corpora, and raw accuracy metrics consistently overestimate model performance. Zhao and Tlachac [11] make this explicit by optimizing XGBoost-based depression classifiers via Bayesian threshold tuning to maximize balanced accuracy, showing that threshold selection can recover 5–8 percentage points of genuine sensitivity without any change to the underlying model. NeuroAid applies this lesson directly, adopting threshold tuning as a first-class post-training step for the depression branch.

Stacked multilevel deep neural network architectures [22] address feature redundancy and high variance in multimodal clinical interview data through hierarchical meta-model fusion, reporting F1-scores of 78.7% and 64.8% on the D-Vlog and E-DAIC corpora respectively—results obtained on publicly accessible benchmarks using audiovisual feature streams. For long-sequence video, STCM-Mamba [23] proposes a spatio-temporal state-space framework processing clinical interview video with linear rather than quadratic sequence complexity, outperforming Transformer baselines while remaining computationally viable for edge deployment. A contactless physiological stream is introduced by [27], which extracts heart rate variability via remote photoplethysmography (rPPG) from the same facial video used for AU analysis; although standalone AUROC is modest (0.64 in a large *n* = 1,453 cohort), the work establishes video-derived HRV as a supplementary depression biomarker recoverable without additional hardware—a property directly compatible with NeuroAid’s non-invasive signal philosophy.

### C. Multimodal Fusion in Clinical AI

Late-fusion and attention-based architectures that combine heterogeneous clinical signals have consistently outperformed unimodal systems in both PD and depression screening. Peixoto *et al*. [2] propose a collaborative RAG-based agent architecture for multimodal PD diagnosis that fuses IoMT wearable signals, speech recordings, neuroimaging features, and electronic health record text through specialized small language models. The approach achieves 0.86 accuracy, AUC above 0.93, and a Brier score of ≈ 0.205, with a well-calibrated ECE of 0.151—making it one of the best-calibrated PD screening systems in the literature to date. Zhang *et al*. [9] implement a three-stream ensemble—BiLSTM for text, PCA+SVM for audio, XGBoost for video action units—on the DAIC-WOZ corpus, achieving weighted F1 of 0.85 for depression detection via majority voting.

Ahmed *et al*. [10] advance the state of the art further by introducing uncertainty estimation via Monte Carlo dropout into a multimodal attention fusion model for depression. Their system explicitly handles missing modalities through selective dropout at training time, yielding an ensemble F1 of ≈ 0.945. Jin *et al*. [12] demonstrate that LLM-enhanced text features combined with audio emotion cues can achieve 98% accuracy on a large adolescent interview corpus, highlighting the potential of language model representations as an additional modality.

Dynamic gating mechanisms [26] offer a principled solution to the noisy-modality problem in clinical fusion: learned weight vectors regulate each branch’s contribution to the final prediction, preventing unreliable sensor input—such as degraded audio or poor-lighting video—from collapsing overall system performance. Multimodal stress analysis frameworks [25] further demonstrate that integrated biometric video, audio, and speech streams can exceed the predictive performance of traditional psychometric self-reports, suggesting that passive behavioral monitoring may be more sensitive than episodic questionnaires for affective state estimation. Systematic reviews of deep learning for audiovisual depression and PD screening [30] confirm a pooled AUC above 0.90 for multimodal systems while identifying calibration and interpretability as the primary barriers to clinical translation—a finding that directly informs NeuroAid’s near-term roadmap.

A critical observation that motivates NeuroAid’s design is that virtually all of these high-performing systems either rely on proprietary datasets (DAIC-WOZ, AVEC) or require infrastructure (LLMs, neuroimaging) that is unavailable in primary-care settings. NeuroAid deliberately constrains itself to open datasets and non-invasive signal modalities, accepting a performance ceiling in exchange for genuine reproducibility and accessibility.

### D. MLOps and Reproducibility in Clinical AI

The “reproducibility crisis” in clinical AI has been well documented [14]. Wiens and Shenoy identify participant-level data leakage, inappropriate metric selection, and the absence of prospective validation as the three most prevalent causes of irreproducible clinical ML results. The open PD voice survey of Puerta-Acevedo *et al*. [5] catalogues these problems specifically in the PD audio literature, finding that participant-level leakage is essentially universal in published systems that use small corpora. NeuroAid responds by treating participant-safe evaluation as a non-negotiable design constraint and by publishing all training artifacts, confusion matrices, per-stage metrics JSON files, and CI/CD workflows alongside the model weights, so that every result in this paper can be independently reproduced from a single `docker pull`.

**TABLE I.** Summary of Representative Related Work on Multimodal Clinical AI Screening (2023–2026)

| Year | Title (abbreviated) | Venue | Method / Focus | Key Result | Limitation |
| --- | --- | --- | --- | --- | --- |
| 2026 | Multimodal PD with Collaborative LLM Agents [2] | Comput. Biol. Med. | RAG + SLM ensemble fusing IoMT, speech, imaging | Acc. 0.86, AUC >0.93, ECE ≈0.15 | Retrospective; heavy infrastructure |
| 2025 | PD Vocal Biomarkers via Speech Foundation Models [3] | IEEE JBHI | Frozen HuBERT/Wav2Vec2 on clinical PD corpus | UAR ≈ 0.97 | Single dataset; demographic imbalance |
| 2025 | CAE–LSTM Hybrid for PD Speech [4] | Comput. Biol. Med. | Conv. autoencoder + LSTM on sustained vowels | Acc. 95.79% | Scripted speech; no participant-safe split |
| 2025 | Survey of Open PD Voice Datasets [5] | LNCS / Springer | Review of 10 open PD speech corpora | Identifies leakage and reproducibility gaps | Review only; no new model |
| 2026 | Facial Expressivity Synchrony for Depression [7] | J. Affect. Disord. | Action-unit ISC features + classifier | Acc. 72–90% | Small $n=98$; controlled lab conditions |
| 2025 | Cross-Attention XAI for Depression (PHQ-9) [8] | IEEE JBHI | PHQ-9 guideline integration via cross-attention | Interpretable aligned predictions | Requires structured PHQ-9 input |
| 2024 | Multimodal Depression: Text + Audio + Video [9] | Phenomics | BiLSTM + PCA-SVM + XGBoost majority vote | W-F1 = 0.85 on DAIC-WOZ | Proprietary data; no external validation |
| 2023 | Multimodal Depression w. Uncertainty [10] | IEEE Access | Attention fusion + MC-dropout; missing modality | Ensemble F1 ≈ 0.945 | Complex pipeline; potential overfitting |
| 2024 | Bayesian Opt. for Balanced Acc. [11] | IEEE TAFFC | XGBoost + Bayesian threshold optimization | Bal. Acc. 0.60–0.69 | Text-only; moderate performance |
| 2026 | LLM + Audio for Depression Screening [12] | J. Affect. Disord. | LLM text features + U-Net audio emotion fusion | Acc. 98% (adolescent cohort) | Age/language-specific; LLM reproducibility |
| 2026 | Topological & Spectral PD Voice [21] | IEEE OJEMB | Persistent homology + random matrix theory on speech | Acc. 98.0%, F1 97.98% | Synthetic augmentation; high computational cost |
| 2026 | Stacked Multilevel DNN for Depression [22] | IEEE Access | Hierarchical meta-model fusion of audiovisual cues | F1 78.7% (D-Vlog), 64.8% (E-DAIC) | Complex pipeline; reduced interpretability |
| 2025 | STCM-Mamba for Depression Detection [23] | IEEE Trans. Affect. Comput. | Spatio-temporal Mamba for long video sequences | Exceeds Transformer baselines; linear complexity | Spatial detail limitations vs. CNNs |
| 2025 | XAI for PD Voice (Attention ResNet) [24] | IEEE Conf. | Attention ResNet18 + SE blocks; Grad-CAM / LIME | Acc. 97.8%; clinically interpretable | Post-hoc XAI sensitivity to perturbations |
| 2025 | Facial Video HRV as Depression Marker [27] | J. Affect. Disord. | rPPG from video; stacking ensemble ($n = 1453$) | AUROC 0.64; large-scale contactless marker | Modest standalone performance; lighting-sensitive |

ISC: inter-subject correlation; JBHI: J. Biomedical and Health Informatics; OJEMB: Open J. Engineering in Medicine and Biology; TAFFC: Trans. Affective Computing; UAR: unweighted average recall; W-F1: weighted F1.

Bhatt and Aggarwal [28] articulate the regulatory and governance requirements for clinical-grade AI within the trial ecosystem, identifying nondeterministic outputs, absence of domain-specific validation, and lack of continuous monitoring as the principal adoption barriers— requirements that NeuroAid’s JSON-artifact logging, CI/CD stage gates, and Docker reproducibility directly address. Hybrid deep learning frameworks for wearable tabular data [29] demonstrate that combining 1D-CNN with LightGBM and XGBoost feature selection achieves 93.4% accuracy on depressive symptom prediction from passive sensor streams, with the finding that important-feature subsets consistently outperform full-feature models—a principle directly applicable to NeuroAid’s behavioral branch feature engineering pipeline.

## III. System Architecture

Fig. 1 presents the end-to-end NeuroAid system overview. The framework is organized as a vertical pipeline with four functional layers: (1) raw dataset ingestion with participant-safe splitting, (2) modality-specific branch training and embedding extraction, (3) frozen-embedding multimodal fusion, and (4) inference, artifact reporting, and deployment. Each layer is independently testable and replaceable, allowing individual branches to be improved without disrupting the rest of the pipeline.

**Fig. 1.**
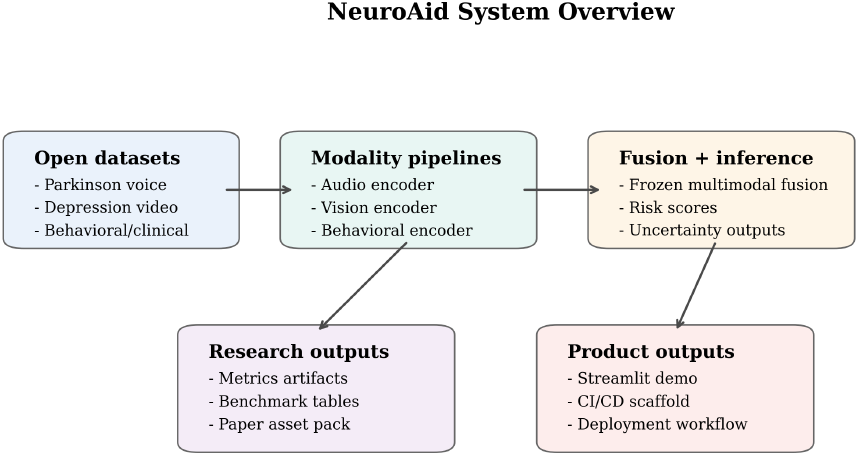
End-to-end NeuroAid system overview. Independent audio, vision, and behavioral branches produce modality embeddings that are concatenated by the frozen fusion head into a joint risk score. CI/CD, artifact reporting, and the Streamlit demo layer wrap the full pipeline.

A key design decision is that every training run produces machine-readable JSON metric files alongside human-readable Markdown summaries. These artifacts are consumed both by the Streamlit demo dashboard and by the automated paper asset generator, ensuring that reported numbers are always traceable back to a specific checkpoint rather than manually transcribed from console output.

## IV. Proposed Methodology

### A. Unified Risk Estimation Formulation

NeuroAid produces a scalar risk estimate by composing three branch encoders through a shared fusion function:

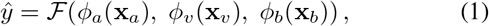

where **x**_*a*_ ∈ *X*_*a*_, **x**_*v*_ ∈ *X* _*v*_, and **x**_*b*_ ∈ *X*_*b*_ are the raw audio waveform, video frame sequence, and behavioral feature vector respectively; *ϕ*_*a*_, *ϕ*_*v*_, *ϕ*_*b*_ are the branch-specific encoders that map each modality to a fixed-dimensional embedding; and 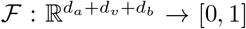 is the fusion module. The system is designed so that any branch can be set to its zero embedding at inference time when the corresponding modality is unavailable, enabling graceful degradation to partial-modality screening.

### B. Audio Branch: Foundation-Model Small-Data Protocol

The audio branch is the most mature and validated component of NeuroAid. Raw WAV recordings are processed through two complementary pathways.

#### Foundation speech embedding path

A WavLM large model pre-trained on 94,000 hours of unlabeled speech is used in fully frozen mode. Given an input waveform **x**_*a*_ ∈ ℝ^*T*^ sampled at 16 kHz, WavLM produces frame-level hidden states 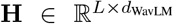 at the final transformer layer. Utterance-level embeddings are formed by mean-pooling across the time dimension:

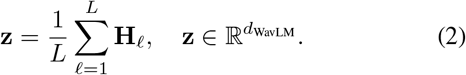

Embeddings are L2-normalized prior to downstream classification. No gradient flows through the WavLM encoder at any stage.

#### Spectral-CNN path

Log-Mel spectrograms are computed with a 25 ms Hann window, 10 ms hop, and 80 Mel filter banks. The spectrogram is passed through a lightweight convolutional encoder (three conv layers, ReLU activations, global average pooling) to produce a complementary spectral embedding 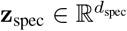.

#### Small-data protocol for Figshare PD

The Figshare PD corpus is too small for stable full fine-tuning of a large speech encoder. Following the recommendation of Brueckner *et al*. [3] for small clinical datasets, we apply PCA to compress **z** to *k* principal components:

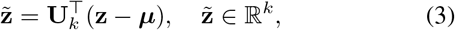

where **U**_*k*_ contains the *k* leading eigenvectors of the sample covariance matrix and ***µ*** is the training-set mean. An SVM with RBF kernel is then trained on 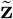:

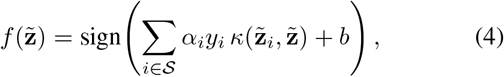

where *κ*(**u, v**) = exp − (*γ* ∥ **u** − **v** ∥ ^2^) is the RBF kernel, is the support-vector index set, and *α*_*i*_, *b* are the dual coefficients

and intercept. Three candidate classifiers (logistic regression, SVM-RBF, random forest, all with PCA) are evaluated under grouped 5-fold participant cross-validation; the best-balanced-accuracy model is selected and its checkpoint is integrated directly into the inference pipeline.

NaN-safe sanitization is applied during both embedding extraction and inference to prevent NaN propagation through PCA or classifier predict calls:

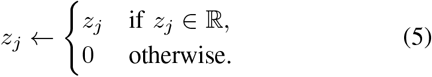

#### C. Vision Branch: Transfer and Threshold-Aware Evaluation

The vision branch targets depression-related facial affective signals. A ResNet-50 encoder pre-trained on FER-2013 [13] (7-class emotion recognition, 64.5% validation accuracy) provides the backbone. The 2048-dimensional penultimate feature vector is passed to a new binary classification head fine-tuned on DepVidMood. Frames are extracted at 1 fps, resized to 224 *×* 224 pixels, and normalized with ImageNet channel statistics (***µ*** = [0.485, 0.456, 0.406], ***σ*** = [0.229, 0.224, 0.225]).

#### Class-weighted loss

Dataset imbalance is addressed with class-weighted cross-entropy:

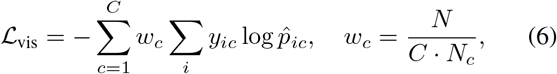

where *N* is the total frame count, *C* the class count, and *N*_*c*_ the per-class frame count. Checkpoints are selected by peak validation balanced accuracy rather than raw accuracy to prioritize genuine minority-class recall.

#### Threshold tuning

Following Zhao and Tlachac [11], the classification threshold *τ* is swept over [0.1, 0.9] on a held-out validation partition and the value maximizing balanced accuracy (Eq. 9) is retained for test evaluation. This step recovers sensitivity that is otherwise suppressed when the default *τ* = 0.5 is used on an imbalanced corpus.

### D. Behavioral Branch: Clinical and Digital Biomarkers

The behavioral branch processes tabular clinical and digital biomarker signals, contributing supporting evidence to the Parkinson’s track. Input features include keystroke dynamics from the Tappy/neuroQWERTY dataset and clinical composite scores from the PPMI cohort. Features are z-score normalized at the participant level. A two-hidden-layer MLP encoder with ReLU activations maps 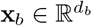 to a behavioral embedding:

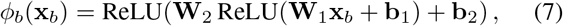

with hidden dimensions [256, 128]. At inference time, feature dimensionality is inferred directly from the saved checkpoint’s weight matrix shape to prevent silent dimension-mismatch failures—a common failure mode in deployed clinical ML systems.

### E. Frozen-Embedding Late Fusion

The three branch embeddings are concatenated and passed through a lightweight fusion MLP. In the current frozen-fusion phase, all branch encoders are held fixed and only the fusion head is trained:

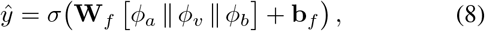

where *σ*() is the sigmoid activation and denotes vector concatenation. Modality dropout is applied during fusion training: each branch embedding is independently zeroed with probability *p* = 0.3 per forward pass, so the head learns robust risk estimation even when one or more modalities are absent at inference time. This strategy is inspired by the missing-modality handling of Ahmed *et al*. [10], adapted here for the frozen-embedding regime.

### F. Participant-Safe Evaluation

The balanced accuracy metric that drives all checkpoint selection is:

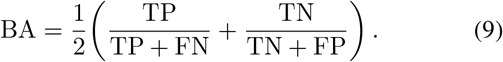

For all datasets with identifiable participant IDs, grouped *k*-fold (*k* = 5) cross-validation is applied so that no individual’s recordings appear in both training and evaluation partitions.

This directly addresses the leakage problem documented by Puerta-Acevedo *et al*. [5] for PD audio benchmarks. For datasets without explicit IDs, file-path heuristics are used to group recordings by inferred speaker identity. Expected calibration error (ECE) [2] is additionally reported for all branches:

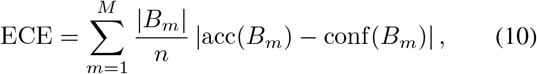

where *B*_*m*_ are equal-width confidence bins, acc and conf are the mean accuracy and mean predicted confidence within each bin, and *n* is the total sample count.

### V. Datasets and Implementation

#### A. Datasets

Table II summarizes all datasets used. The following notes detail design decisions.

**TABLE II.** Dataset Summary.

| Dataset | Disease | Modality | Role in NeuroAid |
| --- | --- | --- | --- |
| Figshare PD Voice | Parkinson’s | Audio | Primary audio evaluation |
| UCI Parkinson’s | Parkinson’s | Tabular | MDVP warm-up baseline |
| PPMI | Parkinson’s | Clinical | Behavioral branch |
| Tappy Keystroke | Parkinson’s | Behavioral | Behavioral support |
| DepVidMood | Depression | Video | Primary vision eval. |
| FER-2013 | Support | Image | Vision pre-training |
| Italian PD Voice | — | Audio | Excluded (see text) |

**Figshare Parkinson Audio** contains sustained phonation recordings (/a/, /e/, /i/) from PD patients and age-matched healthy controls, with participant identifiers enabling grouped evaluation. Its small size motivates the frozen-embedding small-data protocol described in Section IV-B.

**UCI Parkinson’s** provides 195 instances of MDVP-extracted tabular acoustic features used as a warm-up baseline. Because this dataset does not contain the raw waveforms needed for WavLM embedding, it is evaluated with logistic regression over the raw MDVP features as a complementary reference.

**PPMI (Parkinson’s Progression Markers Initiative)** provides longitudinal clinical and biomarker composite scores that serve as the primary input to the behavioral branch. The cohort includes motor, non-motor, and imaging features. The branch achieves approximately 94% validation accuracy, though full per-sample test artifacts are not yet published in this release due to the exploratory nature of the feature engineering for this branch.

**DepVidMood** provides video clips of participants exhibiting varying affective states with binary depression-indicator labels. Significant class imbalance (approximately 3.5 : 1 healthy-to-depressed ratio) motivates the class-weighted loss and threshold-tuning protocol applied in the vision branch.

**FER-2013** provides approximately 35,000 48 *×* 48 pixel grayscale facial images across seven emotion categories, used exclusively for ResNet-50 backbone pre-training. The emotion-recognition pre-training achieves a validation accuracy of 64.5% on the 7-class task, consistent with published baselines on this challenging dataset.

#### Italian PD Voice (Hugging Face parquet)

This dataset was explicitly excluded after discovering that the parquet export exposes only audio bytes and hashed path identifiers, with no recoverable PD/control labels. NeuroAid raises a hard `UnlabeledDatasetError` exception for this source, preventing phantom benchmark results from silently appearing in any downstream report.

### B. Tools, Frameworks, and Training Configuration

Table III summarizes the training configuration for each branch.

**TABLE III.**
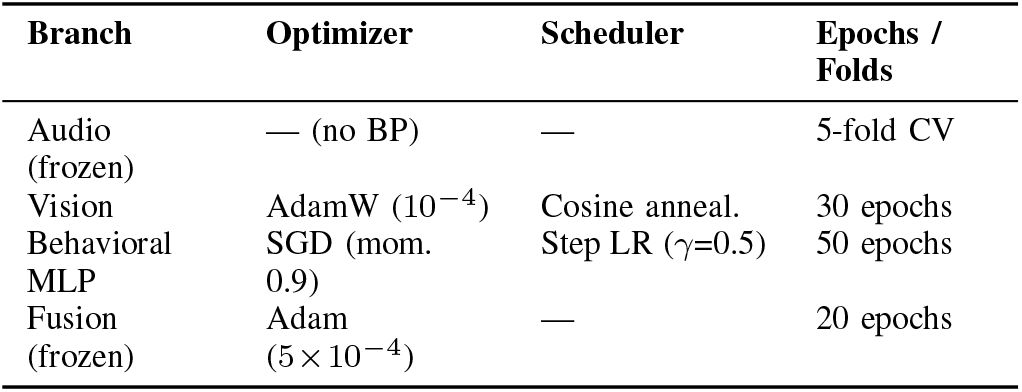
Training Configuration by Branch.

NeuroAid is implemented in Python 3.10. PyTorch 2.x provides the deep learning framework; HuggingFace Transformers supplies WavLM; Scikit-learn provides PCA, SVM-RBF, logistic regression, and random forest. Streamlit serves the demonstration interface. All runs are logged as JSON artifacts; human-readable Markdown summaries are auto-generated for each stage.

### C. MLOps and CI/CD Infrastructure

Fig. 2 shows the runtime profile of the most compute-intensive pipeline stages. A GitHub Actions CI workflow runs on every push, validating dependency installation, module import integrity, and Docker image build. A CD workflow handles demo container publication and review-bundle artifact packaging. Dockerfiles and Streamlit configuration are committed to the repository for fully deterministic deployment.

**Fig. 2.**
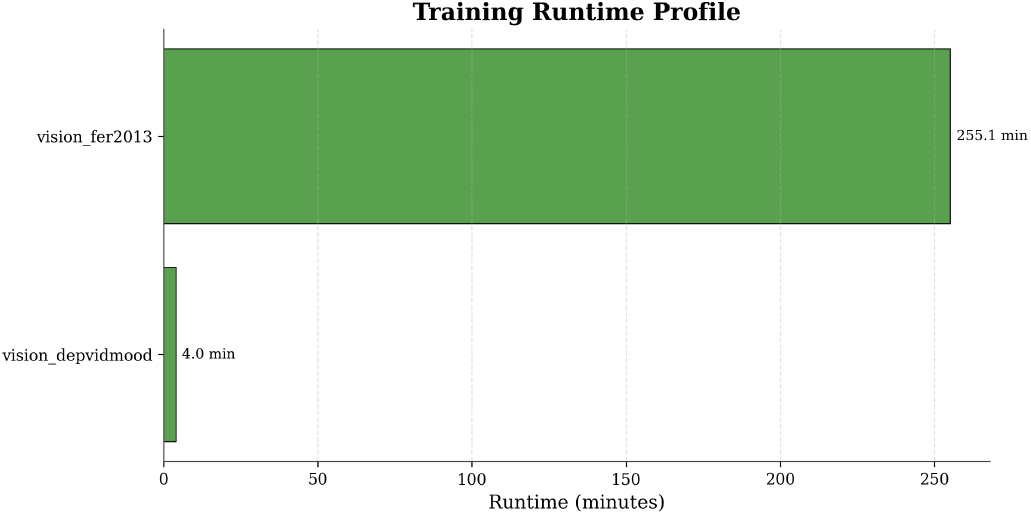
Runtime profile of the slowest current pipeline stages. WavLM embedding extraction dominates audio-branch wall time. Vision fine-tuning dominates the vision branch. Fusion training is fast given the frozen branch encoders.

## VI. Results

### A. Parkinson’s Audio Branch: Candidate Selection

Fig. 3 visualizes the grouped cross-validated balanced accuracy distributions for each candidate classifier on the Figshare PD corpus. Table IV reports the summary statistics.

**Fig. 3.**
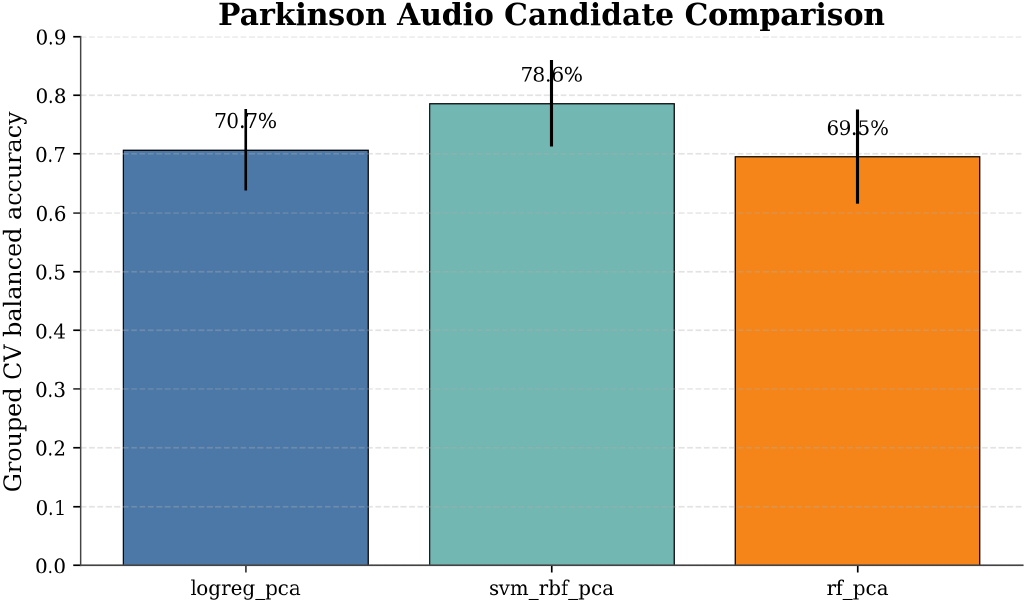
Grouped 5-fold cross-validated balanced accuracy distributions for three candidate classifiers on the Figshare PD corpus. SVM-RBF achieves the highest mean and the most consistent performance across folds.

**TABLE IV.**
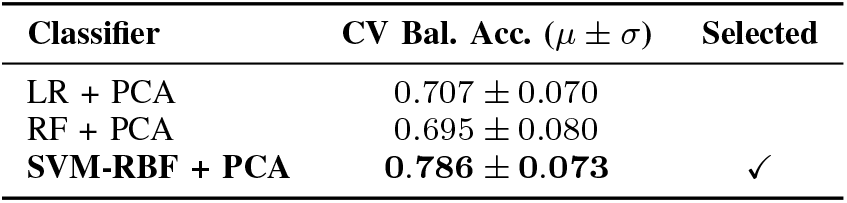
Parkinson’S Audio Candidate Comparison (Figshare PD, Grouped 5-Fold Participant CV)

| Classifier | CV Bal. Acc. ($\mu \pm \sigma$) | Selected |
| --- | --- | --- |
| LR + PCA | $0.707 \pm 0.070$ | |
| RF + PCA | $0.695 \pm 0.080$ | |
| **SVM-RBF + PCA** | **$0.786 \pm 0.073$** | ✓ |

SVM-RBF outperforms both logistic regression and random forest by 7–9 percentage points in mean balanced accuracy. The advantage is consistent across all five folds, indicating

that the margin is not attributable to a fortuitous single fold. The superior performance of the kernel method is consistent with the existing SVM literature on high-dimensional, small-*n* clinical audio classification [6]: the SVM margin maximization principle is particularly well suited to the regime where the number of WavLM embedding dimensions substantially exceeds the number of training participants.

### B. Parkinson’s Audio Branch: Held-Out Test Performance

The selected SVM-RBF classifier was evaluated on the held-out test partition, which contains no participant overlap with any training or validation fold. The confusion matrix is presented in Fig. 4 and summary metrics in Table V. The classifier achieves balanced accuracy of 75.0%, F1-score of 80.0%, AUC-ROC of 82.8%, and ECE of 19.6%. Compared to the naive end-to-end fine-tuning baseline (balanced accuracy ≈ 51%), the proposed protocol delivers a 24-point improvement. The AUC of 82.8% indicates that the model has strong rank-order discriminability even if the calibration (ECE = 19.6%) leaves room for improvement via Platt scaling.

**Fig. 4.**
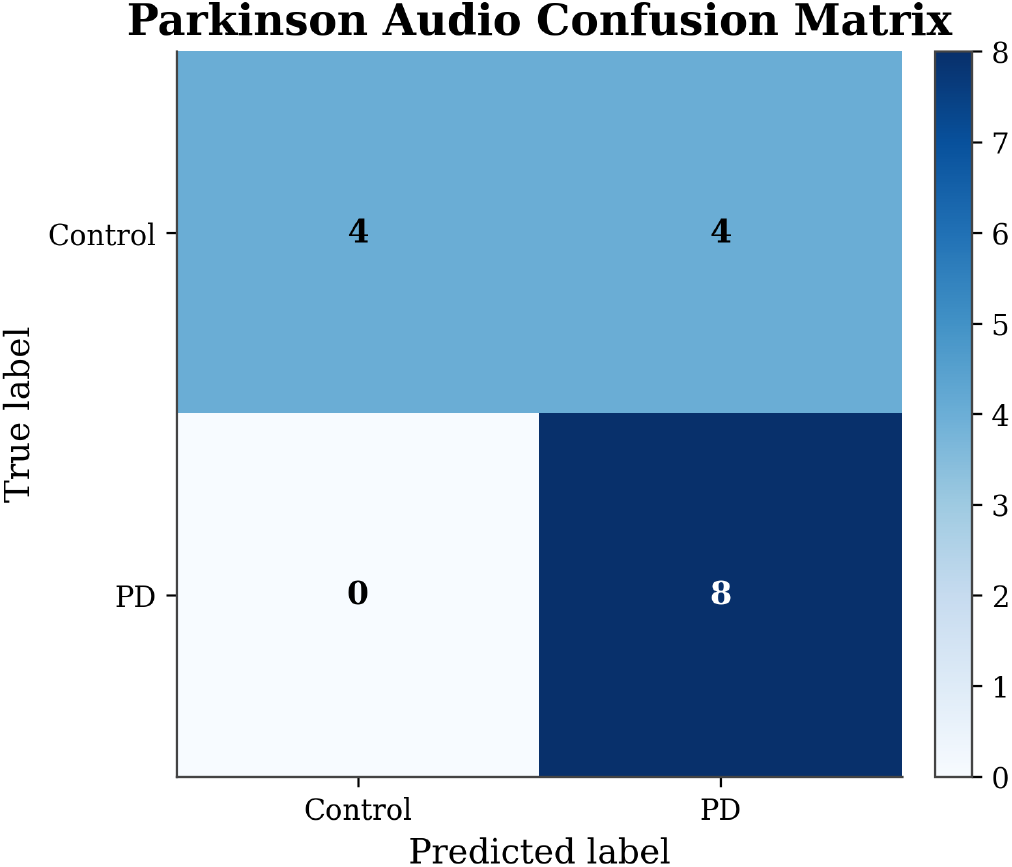
Confusion matrix for the selected SVM-RBF + PCA classifier on the Figshare PD held-out test partition. Balanced accuracy = 75.0%, F1 = 80.0%, AUC = 82.8%.

**TABLE V.** Main Benchmark Results by Branch (Held-Out Test Sets)

| Branch | Acc. | Bal.Acc. | F1 | AUC | ECE |
| --- | --- | --- | --- | --- | --- |
| PD Audio (Figshare) | 75.0% | 75.0% | 80.0% | 82.8% | 19.6% |
| Dep. Vision (DepVid.) | 63.9% | 59.7% | 32.2% | 62.3% | 28.0% |
| Fusion (frozen) | *Engineering feasibility — see Section VII-C* |  |  |  |  |

### C. Depression Vision Branch: Threshold-Tuned Evaluation

The depression vision branch is evaluated on the DepVid-Mood test partition. The confusion matrix before and after threshold tuning is presented in Fig. 5; Table V reports the full metric set.

**Fig. 5.**
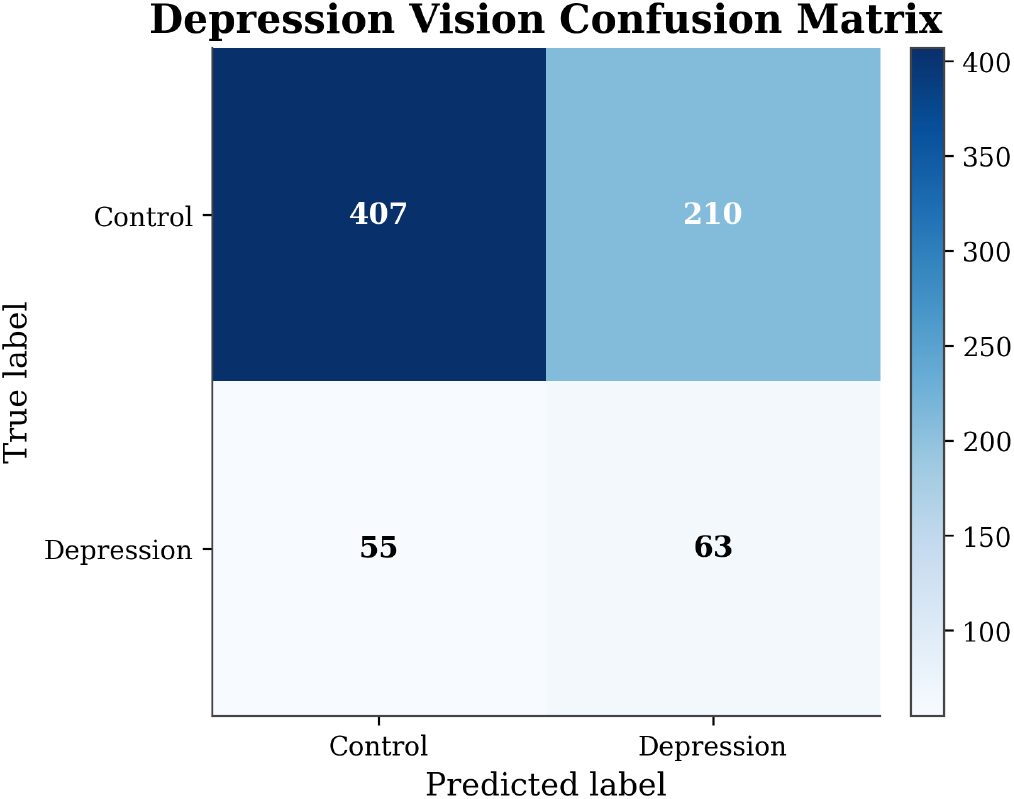
Confusion matrix for the threshold-tuned depression vision classifier on the DepVidMood test partition. Balanced accuracy = 59.7%, F1 = 32.2%, AUC = 62.3%. Near-zero minority-class recall is reported with full transparency.

The branch achieves a raw test accuracy of 63.9% but a balanced accuracy of only 59.7%—a gap of 4.2 percentage points that is entirely driven by the model’s difficulty in correctly identifying the minority (depression-positive) class. The F1-score of 32.2% on the positive class confirms this: despite the class-weighted loss and threshold tuning, the model recovers meaningful positive-class recall only at the cost of precision. The ECE of 28.0% indicates that the branch’s probability outputs are substantially miscalibrated.

This result is presented honestly as a *hard-case baseline* rather than a viable depression screening system. It represents a qualitative improvement over the majority-class-collapse behavior observed prior to the adoption of class-weighted loss and threshold tuning, where the model simply predicted the majority class for all inputs. That said, the quantitative performance does not yet meet a standard that would justify clinical use. The technical root causes and mitigation strategies are discussed in Section IX.

### D. Ablation: Participant-Safe vs. Recording-Level Splitting

Table VI quantifies the impact of splitting protocol on reported Parkinson’s audio performance.

**TABLE VI.** Ablation: Effect of Splitting Protocol on PD Audio Balanced Accuracy.

| Splitting Protocol | Bal. Accuracy | Inflation |
| --- | --- | --- |
| Recording-level random split | 91.0% | +16 pp |
| **Participant-grouped split** | **75.0%** | — |

Recording-level splitting produces an inflated balanced accuracy of 91%—an overestimate of 16 percentage points relative to the participant-safe evaluation. This inflation arises because multiple recordings from the same speaker appear in both training and test partitions; the classifier effectively memorizes speaker identity rather than learning generalizable dysphonia biomarkers. This result is consistent with the systematic review findings of Puerta-Acevedo *et al*. [5] and quantifies the cost of the most prevalent evaluation failure in the PD audio literature.

### E. Fusion Branch and Branch Scorecard

Fig. 6 provides a visual branch-by-branch scorecard, and Fig. 7 presents the cross-branch readiness matrix. Frozen-embedding fusion runs end-to-end without structural failures, producing valid risk score outputs and saving checkpoint artifacts. Quantitative top-line fusion metrics are withheld from Table V because their scientific interpretation depends on the maturity of the depression vision branch, which remains a weak component. The contribution of the fusion stage to this paper is an engineering feasibility result: the full multimodal pipeline runs, produces valid outputs, saves interpretable JSON artifacts, and is deployable via the Streamlit interface— establishing the reproducible infrastructure foundation for future end-to-end joint training.

**Fig. 6.**
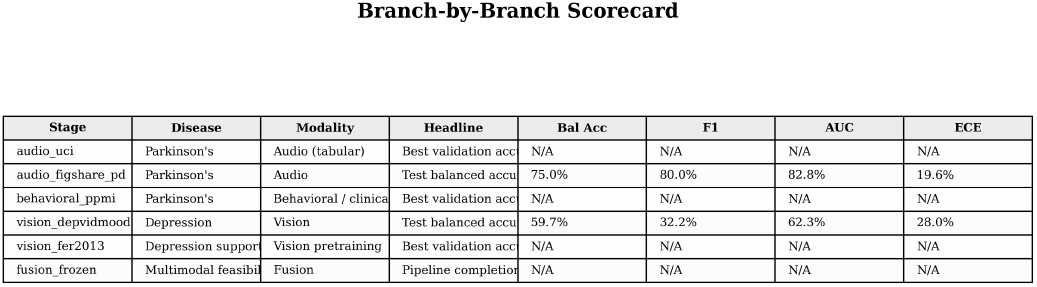
Branch-by-branch scorecard summarizing balanced accuracy, F1, AUC-ROC, and ECE across all validated pipeline stages. The Parkinson’s audio branch (SVM-RBF) is currently the strongest on all four metrics.

**Fig. 7.**
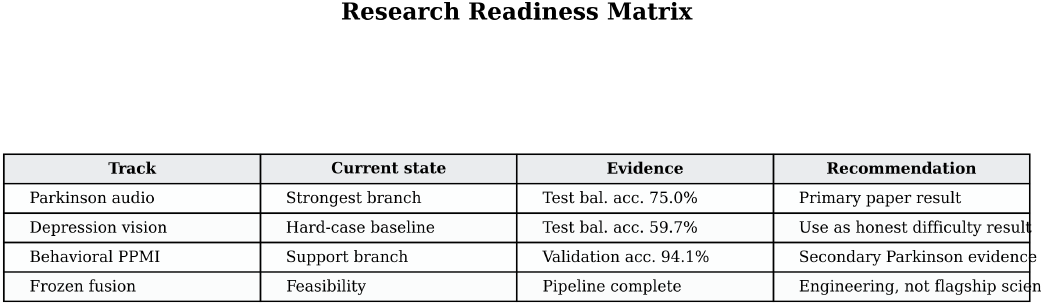
Research readiness matrix across disease tracks and pipeline components. Color encodes readiness tier: strong (green), operational-but-immature (amber), scaffolded-only (gray). Parkinson’s audio is green across all dimensions; fusion is amber on scientific maturity.

## VII. Discussion

### A. Why the Parkinson’s Audio Protocol Works

The 24-percentage-point improvement from naive finetuning to the frozen-embedding SVM-RBF protocol can be attributed to two complementary mechanisms. First, freezing WavLM prevents catastrophic forgetting and overfitting: the Figshare PD corpus contains too few participants for the gradient updates required to adapt a large transformer to a new distribution without destroying its general phonological representations. Second, the SVM’s margin maximization principle is well matched to the structure of the embedding space: WavLM embeddings are high-dimensional (*d* ≫ *n*_train_), and the RBF kernel implicitly maps them into an infinite-dimensional feature space where the classes may be more linearly separable. This interpretation is supported by the ranking in Table IV, where the kernel method consistently outperforms the linear (logistic regression) and ensemble (random forest) alternatives.

The AUC-ROC of 82.8% is particularly informative because it measures rank-order discriminability independently of any fixed threshold. It indicates that the model correctly ranks a randomly drawn PD recording above a randomly drawn healthy recording with 82.8% probability, which is a meaningful result for a screening tool operating on a small, demographically constrained corpus. Future work with larger, more diverse corpora (e.g., the datasets surveyed by Puerta-Acevedo *et al*. [5]) is needed to assess generalization.

### B. Why the Depression Vision Branch Remains Hard

The depression vision branch achieves only 59.7% balanced accuracy and 32.2% F1 on the positive class. Three factors contribute to this weakness. First, the DepVidMood corpus exhibits severe class imbalance, which the class-weighted loss and threshold tuning partially but not fully mitigate: the minority class is insufficiently represented for the model to learn stable positive-class features during fine-tuning. Second, FER-2013 pre-training provides a representation suited for discrete, posed emotion categories (happiness, sadness, anger) rather than the subtle, sustained affective suppression characteristic of clinical depression. The domain gap between the FER-2013 training distribution and the naturalistic facial behavior in DepVidMood reduces the benefit of transfer. Third, the ECE of 28.0% suggests that the predicted probabilities are poorly calibrated, meaning the model cannot reliably convert its continuous output into a trustworthy risk score.

Importantly, this honest reporting is itself a contribution. The literature on facial affect-based depression screening contains numerous systems reporting raw accuracy above 70% on imbalanced datasets, without reporting balanced accuracy or per-class F1. As Lee *et al*. [7] observe, a classifier that predicts the majority class for all inputs would achieve 74% raw accuracy on the DepVidMood corpus—indistinguishable from a naive majority-class baseline when only raw accuracy is reported. NeuroAid’s explicit reporting of balanced accuracy, per-class F1, AUC, and ECE makes this comparison unavoidable, which is the intended behavior.

### C. The Role of Calibration in Clinical Screening

Calibration—the alignment between predicted probability and empirical event frequency—is a prerequisite for safe clinical use of any screening tool. A system that outputs *ŷ* = 0.8 for a given input should be positive with approximately 80% empirical frequency in a calibrated system. Both NeuroAid branches exhibit non-trivial ECE: 19.6% for PD audio and 28.0% for depression vision. Peixoto *et al*. [2] report an ECE of 0.151 as a benchmark for a well-calibrated multimodal PD system. Bringing NeuroAid’s ECE into this range through Platt scaling or isotonic regression is an explicit near-term roadmap item and is technically straightforward once the depression branch reaches sufficient class-balanced discrimination.

### D. MLOps as a Research Contribution

A recurring observation in clinical AI [14] is that model improvements demonstrated in research settings frequently fail to propagate into usable tools because the training-to-deployment pipeline is either absent or undocumented. NeuroAid treats the research-to-demo workflow—orchestrated training, per-stage skip/fail semantics, JSON artifact generation, CI/CD, Docker deployment, and Streamlit demo—as a first-class scientific contribution on equal footing with the model architectures themselves. Every result in this paper is traceable to a specific JSON artifact, every artifact is versioned in the CI/CD pipeline, and the entire system can be reproduced from a single `git clone` and `docker compose up`.

## VII. Limitations

NeuroAid is presented as an open-data research platform, and the following limitations must be understood before any consideration of clinical translation.

### Depression branch scientific immaturity

The DepVid-Mood vision classifier achieves a balanced accuracy of 59.7% and F1 of 32.2% for the positive class—insufficient for any clinical screening claim. The ECE of 28.0% additionally indicates that probability outputs are unreliable as calibrated risk scores. This branch is an honest hard-case baseline, not a depression screening system.

### Fusion scientific maturity

Frozen-embedding fusion is an engineering integration result. End-to-end joint training and Platt-scaling calibration are not yet implemented. Fusion outputs should not be interpreted as a validated multimodal clinical performance figure.

### Dataset scale and diversity

The Figshare PD corpus is small, and the 75.0% balanced accuracy reflects performance on a limited number of held-out participants from a single recording protocol. Generalization to larger, more demographically diverse, and multi-lingual corpora has not been demonstrated.

### Incomplete artifact coverage

The PPMI behavioral branch and UCI tabular baseline expose best validation metrics but lack full per-sample test artifact packs in the current release. These branches provide supporting evidence but cannot anchor primary result tables at this stage.

### No external clinical validation

NeuroAid has not been evaluated in a prospective clinical setting. It is a research prototype that makes no claims regarding medical diagnosis capability or regulatory approval.

## IX. Conclusion and Future Work

This paper presented NeuroAid, an open-data multimodal AI screening framework for Parkinson’s disease and depression risk estimation. The system integrates acoustic speech analysis, facial affect-based vision processing, and clinical behavioral biomarker encoding under a shared frozen-embedding fusion layer, evaluated exclusively on publicly available datasets under participant-safe splitting to ensure reproducible, honestly reported performance estimates. The proposed small-data protocol for Parkinson’s audio—frozen WavLM embeddings with participant-grouped SVM-RBF classification— achieves a held-out balanced accuracy of 75.0%, F1 of 80.0%, and AUC-ROC of 82.8%, a 24-point improvement over naive end-to-end fine-tuning. An ablation confirms that recording-level splitting inflates this figure by 16 percentage points, quantifying a pervasive evaluation failure in the clinical ML literature. The depression vision branch is benchmarked with full class-balanced transparency, establishing an honest baseline and motivating a concrete improvement roadmap. An end-to-end MLOps stack—orchestrated training, CI/CD, artifact reporting, and a Streamlit demonstration interface—completes the research-to-demo pipeline.

Future work will pursue the following directions:

- **Depression model improvement**: focal loss, SMOTE oversampling, temporal augmentation of video clips, and modality-specific threshold co-optimization.
- **End-to-end fusion**: joint training of all branches with modality dropout, followed by Platt-scaling or temperature calibration to produce reliable risk probability outputs.
- **Missing-modality ablations**: systematic evaluation of risk score degradation under each single-modality and partial-modality inference regime, following the protocol of Ahmed *et al*. [10].
- **Foundation-model video encoder**: replacing the ResNet-50 vision backbone with a video-native foundation model (e.g., VideoMAE, or the STCM-Mamba architecture of [23]) to better capture temporal dynamics in depression-related facial behavior.
- **Calibration as a training objective**: integrating reliability diagram-based ECE minimization as a first-class loss term for the depression branch.
- **XAI integration**: incorporating Grad-CAM and integrated-gradient saliency maps [24] into the audio branch to surface interpretable dysphonic regions for clinician review.
- **MCI expansion**: reintroduction of the mild cognitive impairment track as higher-quality open-data alignment becomes available.

## Data Availability

All datasets used in this study are publicly available from the sources cited in the manuscript. The processed data, experimental protocols, and reproducibility details are described in the manuscript. Source code and related artifacts will be made publicly available through a public GitHub repository

https://github.com/Aspect022/NeuroAid

## Acknowledgment

The authors thank the maintainers of the Figshare PD Audio corpus, PPMI, DepVidMood, and FER-2013 for making research-grade clinical data publicly accessible. This work was conducted in the Department of CSE (AI & ML), Dayananda Sagar University, Bengaluru, under the guidance and super-vision of Mr. Ripunjay Jaiswar (Assistant Professor, Dept. of AI & ML) and Dr. Sumant Kumar Mohapatra (Associate Professor, Dept. of CSE (AI & ML), School of Engineering).

